# Hidden Harm: Quantifying occupational injury under-reporting in veterinary workplaces through modified capture-recapture analysis

**DOI:** 10.1101/2025.11.18.25340469

**Authors:** John S.P. Tulloch, Martin Whiting, Rebecca Jackson, Imogen Schofield

## Abstract

**Introduction:** Occupational injuries are prevalent within the veterinary sector, though their true extent is unknown as evidence indicates widespread under-reporting of injuries. This study aimed to: assess injury under-reporting across roles in UK veterinary practices; and audit the type, frequency, and outcomes of workplace injuries within a large veterinary organisation.

**Methods:** A retrospective audit was conducted on a large veterinary organisation’s accident reporting system in 2022. Under-reporting was estimated using a modified capture-recapture method, comparing audit records with self-reported injury data from a cross-sectional staff survey stratified by role and employment figures. Audit data were descriptively analysed and compared with survey data.

**Results:** The overall injury under-reporting rate was 68.9%; for every 100 workplaces injuries, 69 went un-reported. Levels of under-reporting were higher in companion animal practices (70.0%) than large animal practices (56.4%). Common causes of injury of companion animal staff included; clinical examination (28.2%); falls, slips and trips (11.2%); drug administration (10.4%), and needlesticks and surgical sharps injuries (6.1%).

**Limitations:** Survey responses could not be directly linked to audit records due to anonymity, and survey-based prevalence estimates assumed only one injury per person per year, likely underestimating true injury rates.

**Conclusions:** Occupational injury under-reporting is widespread in UK veterinary practices, particularly companion animal practices. Without improving reporting, it will be challenging to establish the true incidence and context of occupational injuries in the veterinary workforce. Strengthening reporting, training, leadership engagement, and visible responses to incidents are key to strengthening safety culture and injury reporting.

**Highlights:**

- For every 100 veterinary workplace injuries, 69 will go unreported
- In clinical roles, companion animal vets had the highest rate of underreporting,78%
- Needlesticks and hazardous exposures more common than expected
- Many preventable injuries occur to practice visitors
- Reporting can improve with training, leadership, and visible incident responses

## Introduction

Occupational injuries are a well-documented part of veterinary workplaces, with hazards such as animal-related trauma, needlestick injuries, and chemical exposure, being ever present (Gabel et al., 2002; Nienhaus et al., 2005; Fritschi et al., 2006; Fowler et al., 2016; Furtado et al., 2024; Johnson and Fritschi, 2024; Tulloch et al., 2025a). These injuries have high rates of hospital attendance, with potentially life changing consequences (Fritschi et al., 2006; Parkin et al., 2018; Tulloch et al., 2023; Johnson and Fritschi, 2024; Voss et al., 2024; Tulloch et al., 2025a, b). The structure of the UK veterinary clinical sector is comprised of a combination of independent private practices and hospitals, corporate groups owning and operating multiple practices in a single structure, and university practices and hospitals. Due to this diverse structure, systematic surveillance and reporting structures for occupational injuries are limited, with injury-under-reporting thought to be widespread (Nienhaus et al., 2005; Furtado et al., 2024).

One major obstacle to understanding and addressing occupational risk in veterinary workplaces is under-reporting. Reasons for under-reporting in the veterinary sector have included; the injury was perceived to be too minor, the injury was an inevitable everyday hazard not worthy of reporting, individuals were too busy or it was too much effort to file a report, they did not know the reporting process, and that reporting would make no difference (Furtado et al., 2024; Tulloch et al., 2025a, b). Consequentially reported injury rates may only be a fraction of true incidents and the relative frequency of different types of events (i.e. dog bites, musculoskeletal injuries) may not be reflective of their true frequency. Thus, it is challenging to identify what are the true occupational risks faced by veterinary staff and what interventions and policies can be developed to mitigate them.

Under-reporting can occur at two levels, individual reporting to their employer, and organisational reporting to a competent authority (in the UK this is the Health and Safety Executive (HSE)). To calculate the degree of organisational under-reporting the capture-recapture methodology has frequently been used where business safety records are matched to competent authorities’ records (Rosenman et al., 2006; Boden and Ozonoff, 2008; Probst et al., 2008; Orellana et al., 2021). Measuring individual under-reporting is more challenging and has relied on surveys of employees (Probst and Estrada, 2010; Lipscomb et al., 2015; Tulloch et al., 2025a). Few have explored under-reporting in the veterinary profession. Surveys of UK veterinary professionals have estimated that 37% of equine veterinarians, 33% of production animal veterinarians, and 44% of mixed practice veterinarians self-declared that they had reported their most recent injuries to their employer’s injury reporting system (Parkin et al., 2018; Tulloch et al., 2025a). In the companion animal sector, veterinarians reported 47% of their most recent injuries, whilst 74% of veterinary nurses reported theirs (Tulloch et al., 2025b). In an Australian university only 4% of veterinary students reported an equine-related injury to the university (Riley et al., 2015).

We propose that utilising a modified capture-recapture analysis we could estimate a more accurate level of individual injury under-reporting. To achieve this, we will utilise two independent data sources relating to injuries in the CVS UK Ltd groups of practices. There are 430 CVS practices in the UK, representing around 8% of the UK market (plc, 2024). The aims of this study were threefold. Firstly, to ascertain levels of injury under-reporting amongst differing roles in UK veterinary practices. Secondly to audit the reporting, type, frequency, and outcomes of workplace injuries within a veterinary organisation. Thirdly, to compare how officially reported audit data differs from previously published self-reported injuries.

## Method

A retrospective, descriptive audit of recorded workplace injuries sustained in 2022 was undertaken using SafetyHub (CVS Group Ltd’s Health and Safety accident reporting portal). This system is utilised by all those employed in CVS’s companion animal, equine, and production animal clinics and hospitals. To estimate the degree of injury under-reporting we used a modified capture-recapture methodology. The first source of injury data was the SafetyHub audit, and the second source of data was the results of a large cross-sectional survey of CVS employees exploring work-related injuries (Furtado et al., 2024; Tulloch et al., 2025a, b). The percentage of people reporting to have been injured in 2022 was extracted from the survey. Both these datasets were stratified by role. Employment numbers for 2022, stratified by job role, was subsequently provided by CVS. The overall, and stratified, under-reporting rates were calculated using the following formulae:

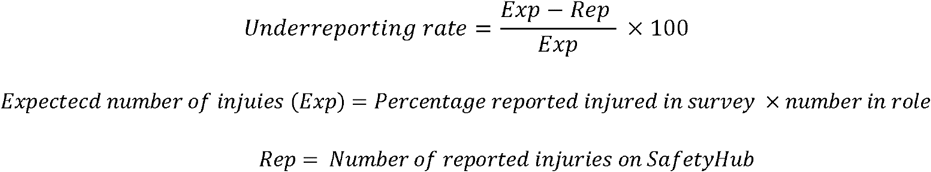

Expectecd number of injuies (Exp) = Percentage reported injured in survey x number in role Rep = Number of reported injuries on SafetyHub

A descriptive analysis of SafetyHub’s data was performed. We stratified this data by practice type (companion animal or large animal), and the role of the person injured (including non-employees). The following contextual information were described; time of day, location of injury within the practice, animal involvement, and mechanism of injury. If enough information was available within the system, further descriptive analysis of the context surrounding the most prevalent injury mechanisms was performed. Consequential information was described including; anatomy injury, type of injury, medical treatment, time off work, whether the injury was RIDDOR (Reporting of Injuries, Diseases and Dangerous Occurrences Regulation 2013) reportable to HSE, and whether immediate action beyond first aid and reporting was taken. In the UK, under RIDDOR, an employer legally must report to the HSE work-related accidents that result in specific injuries (Executive, 2013). These injuries include any that leave them incapacitated for more than seven days, fractures other than to digits, loss of consciousness, among others(Executive, 2013).

Collection and analysis of survey data received ethical approval from the University of Liverpool Veterinary Research Ethics Committee (VREC1256). Data from SafetyHub were originally generated for workplace safety within CVS, not for research. These data were cleaned and anonymised by CVS before sharing with the research team. This project therefore involves the secondary analysis of data and adheres to ethical principles of research integrity, data protection, and responsible use. As such the University of Liverpool Research Ethics team confirmed that no ethical approval was needed.

## Results

There was an overall occupational injury under-reporting rate of 68.9%. For every 100 workplaces injuries that occurred, 69 went un-reported to SafetyHub (Table 1). Levels of under-reporting were significantly higher in companion animal practices (70.0%) than large animal practices (56.4%). The roles with the highest levels of under-reporting were companion animal receptionists, veterinarians and animal care assistants. The lowest under-reporting rates were in companion animal administrators and equine veterinarians and veterinary nurses.

**Table 1.**
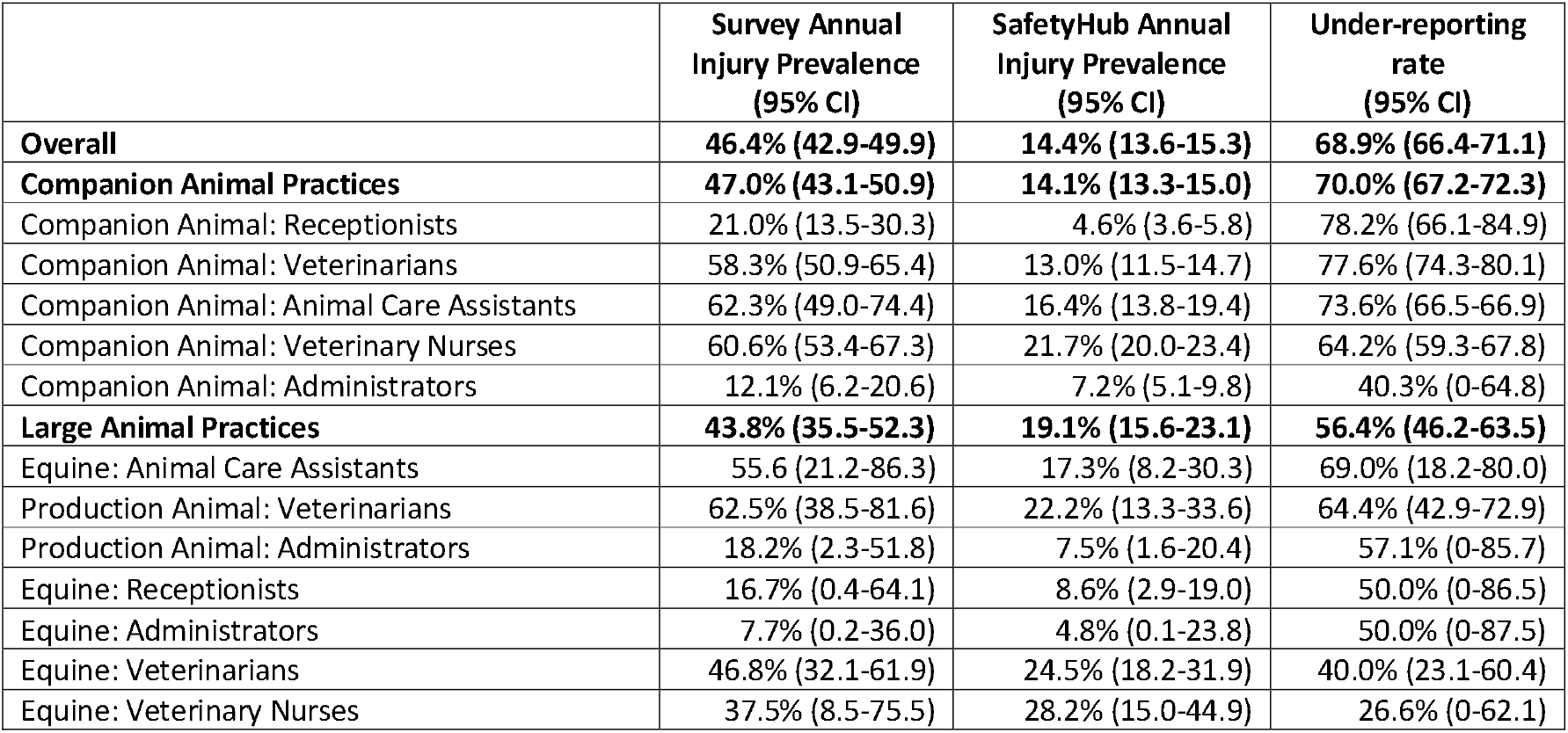
Under-reporting rates of work-related injuries in UK veterinary practices in 2022.

|  | Survey Annual<br>Injury Prevalence<br>(95% CI) | SafetyHub Annual<br>Injury Prevalence<br>(95% CI) | Under-reporting<br>rate<br>(95% CI) |
| --- | --- | --- | --- |
| **Overall** | **46.4% (42.9-49.9)** | **14.4% (13.6-15.3)** | **68.9% (66.4-71.1)** |
| **Companion Animal Practices** | **47.0% (43.1-50.9)** | **14.1% (13.3-15.0)** | **70.0% (67.2-72.3)** |
| Companion Animal: Receptionists | 21.0% (13.5-30.3) | 4.6% (3.6-5.8) | 78.2% (66.1-84.9) |
| Companion Animal: Veterinarians | 58.3% (50.9-65.4) | 13.0% (11.5-14.7) | 77.6% (74.3-80.1) |
| Companion Animal: Animal Care Assistants | 62.3% (49.0-74.4) | 16.4% (13.8-19.4) | 73.6% (66.5-66.9) |
| Companion Animal: Veterinary Nurses | 60.6% (53.4-67.3) | 21.7% (20.0-23.4) | 64.2% (59.3-67.8) |
| Companion Animal: Administrators | 12.1% (6.2-20.6) | 7.2% (5.1-9.8) | 40.3% (0-64.8) |
| **Large Animal Practices** | **43.8% (35.5-52.3)** | **19.1% (15.6-23.1)** | **56.4% (46.2-63.5)** |
| Equine: Animal Care Assistants | 55.6 (21.2-86.3) | 17.3% (8.2-30.3) | 69.0% (18.2-80.0) |
| Production Animal: Veterinarians | 62.5% (38.5-81.6) | 22.2% (13.3-33.6) | 64.4% (42.9-72.9) |
| Production Animal: Administrators | 18.2% (2.3-51.8) | 7.5% (1.6-20.4) | 57.1% (0-85.7) |
| Equine: Receptionists | 16.7% (0.4-64.1) | 8.6% (2.9-19.0) | 50.0% (0-86.5) |
| Equine: Administrators | 7.7% (0.2-36.0) | 4.8% (0.1-23.8) | 50.0% (0-87.5) |
| Equine: Veterinarians | 46.8% (32.1-61.9) | 24.5% (18.2-31.9) | 40.0% (23.1-60.4) |
| Equine: Veterinary Nurses | 37.5% (8.5-75.5) | 28.2% (15.0-44.9) | 26.6% (0-62.1) |

### Companion Animal Practice Employee Reported Incidents

There were 1135 incidents recorded within SafetyHub, 56.8% (n=645) involved employees. Incidents occurred in a bimodal distribution, with peaks occurring between 9-11am and at 3-5pm (Fig1).

**Fig 1.**
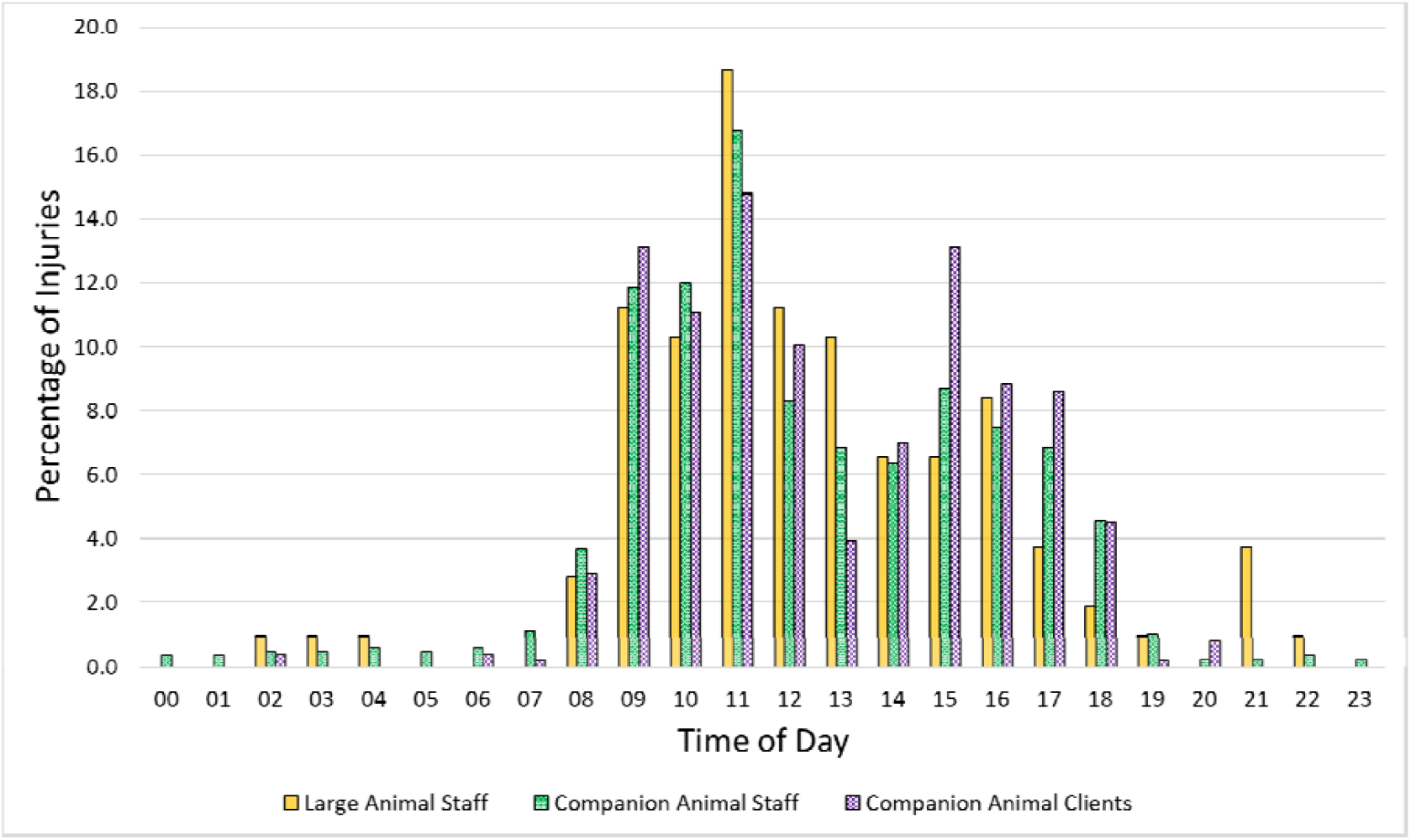
Temporal distribution of reported work-related injuries to staff and clients in UK veterinary practices.

Over half of the reported injuries were to veterinary nurses (37.7%) and veterinarians (23.6%) (Table S1). Veterinary nurses were representative of the national population in terms of gender (95.8% female in SafetyHub vs 96.9% nationally (Surgeons, 2025). Regarding age they were broadly representative with the most prevalent group being 30-39 years old (41.1% vs 40.2% nationally), but they were slightly over-represented in the <30 years old (38.4% vs 29.7% nationally). Veterinary surgeons were less representative with 79% being female (66.8% nationally) and were a younger population with 47.0% being less than 30 years old (25.1% nationally) (Surgeons, 2025).

Injuries primarily occurred in the prep room (32.0%), consultation room (28.1%), and kennels/ward (15.3%) (Table 2). This did vary by role, for example reception staff were injured more in reception (25.6%), and veterinary surgeons were injured more frequently in the consultation room (49.0%) Injuries predominately involved animals (60.3%), with cats and dogs being the most prevalent.

**Table 2.**
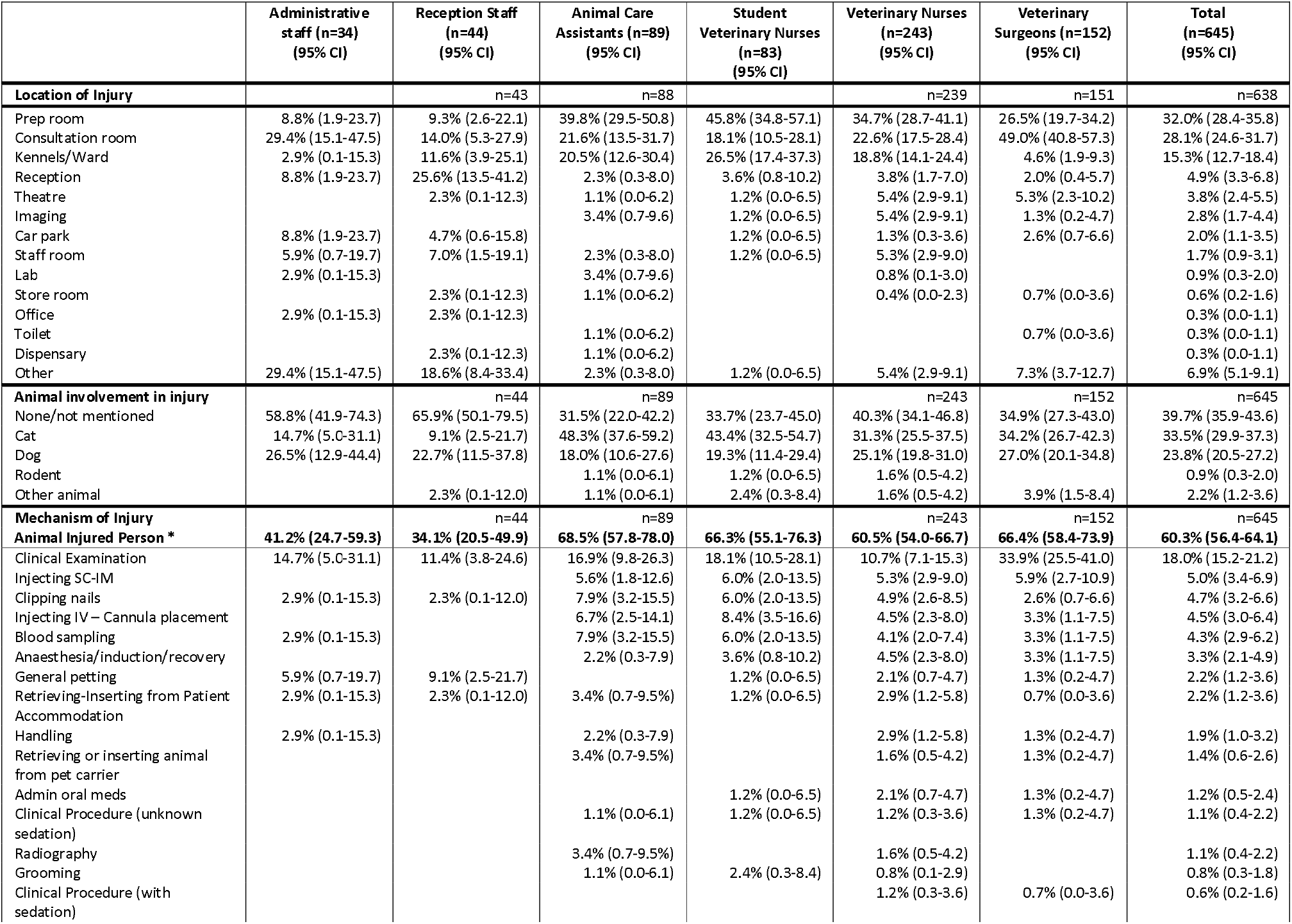
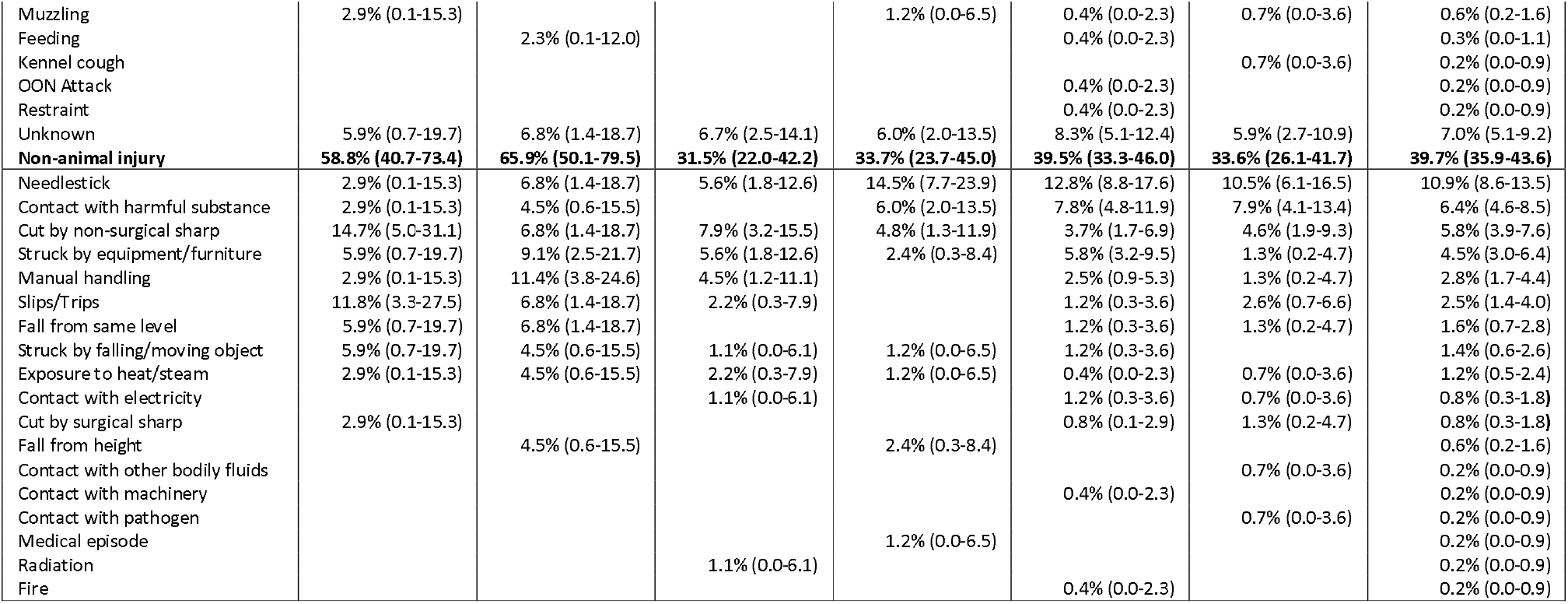
Context of reported work-related injuries in UK companion animal veterinary practices.

The most prevalent mechanisms of injury were a clinical examination (18.0%), needlesticks and surgical sharps (11.7%), drug administration (10.9%), and contact with a harmful substance (6.4%). Only 17.4⍰% (n⍰=⍰20) of injury records from clinical examinations, and none related to drug administration, contained sufficient contextual information for further analysis. Due to this high level of missing data, we did not perform further descriptive analysis. There was limited contextual information about surgical sharps. A third of needlestick injuries occurred whilst performing an injection, however 19.2% occurred through unsafe handling practices (i.e. recapping of needles, passing the device, or sharps being in regular waste (Weese and Jack, 2008)) (Table 3). Additionally, when active ingredients were mentioned, some included sedatives and potential biological hazards. In four cases someone else was injured rather than the person using the needle.

**Table 3.**
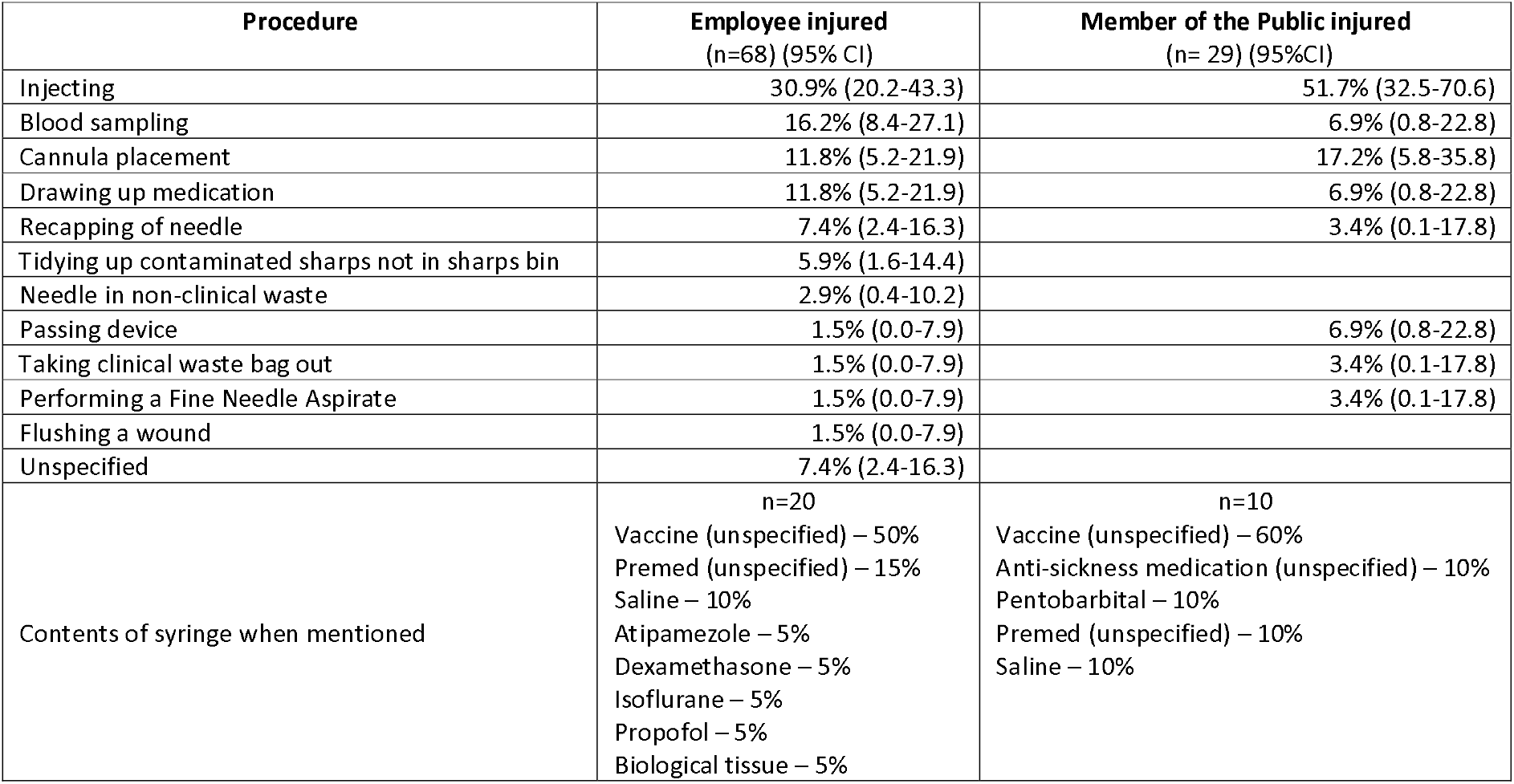
Mechanism of needlestick injuries to companion animal veterinary practices employees and members of the public.

Many harmful substance incidents were reported, with at least 65.9% coming into contact with the IP’s eye (Table S2). Pepper spray was the most prevalent (24.4%), however sedatives as a group made up 29.1% of incidents, and disinfectants another 19.5%.

The main cat-related injuries were puncture wounds (50.2%) and scratches (39.2%), with puncture wounds to the hand (43.8%) and scratches to the hand (27.2%) being the most prevalent injuries (Table S3). Injuries to the head/face/eye made up 4.2% of injuries. The main dog-related injuries were puncture wounds (38.8%), bruises (23.1%), and scratches (22.5%), with puncture wounds to the hand (32.5%) and bruising to the hand (10.6%) being the most prevalent. Fractures made up 1.9% of injuries, whilst 7.0% of injuries were to the head/face/eyes. Dog breed was mentioned in 84.4% of dog-related records. Kennel Club Recognised breeds made up the majority of the incidents (73.3%). Small-sized breeds, as defined by the Kennel Club, were involved in 35.6% of incidents, with large breeds involved in 28.1% (Table S4). The top breeds mentioned were a Staffordshire Bull Terrier (11.1%), Border Collie (9.6%), Labrador Retriever (5.2%), and French Bulldogs (5.2%). Non-animal-related injuries were more diverse, with the main injuries being puncture wounds to the hand (21.2%), cut to the hand (13.4%), and chemical irritation to the eye (10.4%). Fracture made up 0.9% of injuries, whilst 26.9% were to the head/face/eyes, and 0.4% involved a loss of consciousness.

Less than a third of individuals did not receive medical treatment for their injuries (Table 4). Most received first aid, and 5.7% attended a hospital. Over 99% took no time off work for their injuries, and less than 1% were RIDDOR reportable. Actions by practices included; education (topics included; cat scratch disease and removing sharps safely), changes in dangerous drugs stock-taking, review of animal handling, implementing care signs on kennels, and fixing a carpet.

**Table 4.** Consequences of workplace injuries to employees at companion animal veterinary practices.

|  | Administrative<br>staff (n=34)<br>(95%CI) | Reception Staff<br>(n=44)<br>(95%CI) | Animal Care<br>Assistants (n=89)<br>(95%CI) | Student<br>Veterinary Nurse<br>(n=83)<br>(95%CI) | Veterinary Nurses<br>(n=243)<br>(95%CI) | Veterinary Surgeons<br>(n=152)<br>(95%CI) | Total<br>(n=645)<br>(95%CI) |
| --- | --- | --- | --- | --- | --- | --- | --- |
| **Medical Treatment** |  |  |  |  |  |  |  |
| None | 23.5% (10.8-41.2) | 29.5% (16.8-45.2) | 27.0% (18.5-36.9) | 43.4% (32.5-54.7) | 30.9% (25.1-37.1) | 30.3% (23.1-38.2) | 31.3% (27.8-35.1) |
| First aid | 70.6% (52.5-84.9) | 61.4% (45.5-75.6) | 57.3% (46.4-67.7) | 48.2% (37.1-59.4) | 58.8% (52.4-65.1) | 59.2% (51.0-67.1) | 58.1% (54.2-62.0) |
| Primary Care |  | 2.3% (0.1-12.0) | 3.4% (0.7-9.5) | 3.6% (0.8-10.2) | 4.9% (2.6-8.5) | 7.9% (4.1-13.4) | 4.8% (3.3-6.8) |
| Minor Injury Unit |  | 2.3% (0.1-12.0) | 2.2% (0.3-7.9) | 3.6% (0.8-10.2) | 0.8% (0.1-2.9) |  | 1.2% (0.5-2.4) |
| Emergency Department | 5.9% (0.7-19.7) | 4.5% (0.6-15.5) | 10.1% (4.7-18.3) | 1.2% (0.0-6.5) | 4.5% (2.3-8.0) | 2.6% (0.7-6.6) | 4.5% (3.0-6.4) |
| **Time off work** |  |  |  |  |  |  |  |
| None | 100% | 100% | 100% | 98.8% (93.5-100) | 98.8% (96.4-99.7) | 99.3% (96.4-100) | 99.2% (98.2-99.8) |
| <7 days |  |  |  |  | 1.2% (0.3-3.6) | 0.7% (0.0-3.6) | 0.6% (0.2-1.6) |
| 7-14 days |  |  |  | 1.2% (0.0-6.5) |  |  | 0.2% (0.0-0.9) |
| **RIDDOR Reportable** |  |  |  |  |  |  |  |
| Yes |  | 2.3% (0.1-12.0) | 2.2% (0.3-7.9) |  | 1.2% (0.3-3.6) |  | 0.9% (0.3-2.0) |
| No | 100% | 97.7% (88.0-99.9) | 97.8% (92.1-99.7) | 100% | 98.8% (96.4-99.7) | 100% | 99.1% (98.0-99.7) |
| **Immediate Action beyond first aid & report** |  |  |  |  |  |  |  |
| Yes |  |  | 1.1% (0.0-6.1) | 1.2% (0.0-6.5)<sup>1</sup> | 0.8% (0.1-2.9)<sup>2</sup> | 32.0% (0.4-5.7) | 1.1% (0.4-2.2) |
| No | 100% | 100% | 98.9% (93.9-100) | 98.8% (93.5-100) | 99.2% (97.1-99.9) | 98.0% (94.3-99.6) | 98.9% (97.8-99.6) |

### Companion Animal Non-Employee Reported Incidents

There were 490 injuries reported to non-employees within SafetyHub. There was a high degree of missing data for age (98.2%) and gender (94.1%), thus demographics were not explored. Injuries primarily occurred in the consult room (64.3%), however veterinary students were injured in a variety of locations with the prep room being the most prevalent (45.8%). Injuries predominately involved animals (71.8%), with cats and dogs being the most prevalent (Table 5).

**Table 5.**
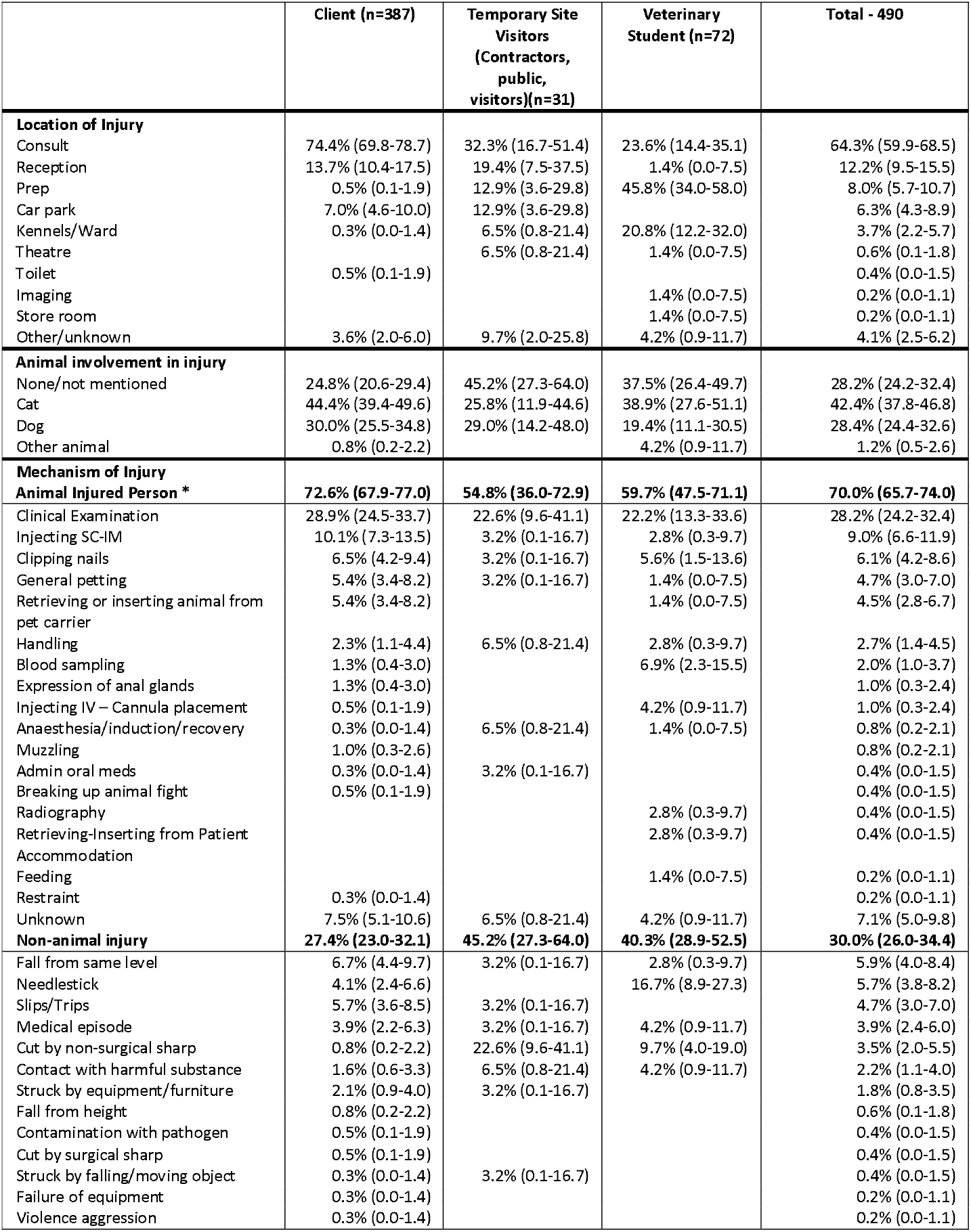
Context of reported injuries to non-employees in UK companion animal practices.

The most prevalent mechanisms of injury were a clinical examination (28.2%); falls, slips and trips (11.2%); drug administration (10.4%), and needlesticks and surgical sharps (6.1%). Medical episodes accounted for 3.9% of injuries. Few records about injuries occurring during clinical examinations or drug administration had enough contextual information for further analysis. Due to this high level of missing data, we did not perform further descriptive analysis. When descriptions of fall, slips and trips (n=55) were described, the most prevalent causes were; 25.5% tripping over animal scales in the waiting room, 21.8% being pulled over by their own dog, 18.2% tripping on an uneven carpark surface, and 14.5% falling down steps into/out of a building. Needlesticks mainly occurred when a veterinarian was administering a drug, and the owner was restraining their animal. (Table 3) Medical episodes predominately were fainting or loss of consciousness events, but also included panic attacks, strokes, and cardiac arrests. The most prevalent resultant injuries were a puncture wound to an arm (28.1%), a scratch to a hand (22.7%), and a cut to a hand (11.7%) (Table S5). Head injuries to the head made up 13.4% of injuries, whilst there were 0.6% fractures, and 1.9% lost consciousness. Around a quarter of individuals did not receive medical treatment for their injuries (Table 6). Most received first aid, and 2.7% attended a hospital, surprisingly some individuals declined medical assistance.

**Table 6.** Consequences of workplace injuries to non-employees at companion animal veterinary practices.

|  | Client (n=387) | Temporary Site Visitor (n=31) | Vet Student (n=72) | Total (n=490) |
| --- | --- | --- | --- | --- |
| **Medical Treatment** |  |  |  |  |
| None | 23.8% (19.6-28.3) | 25.8 (11.9-44.6) | 33.3% (22.7-45.4) | 25.3% (21.5-29.4) |
| Offered medical help, declined/refused | 3.1% (1.6-5.4) |  |  | 2.4% (1.3-4.2) |
| First aid | 58.1% (53.1-63.1) | 64.5% (45.4-80.8) | 55.6% (43.4-67.3) | 58.2% (53.7-62.6) |
| Advised to seek further medical treatment (after 1<sup>st</sup> aid) | 9.8% (7.0-13.2) | 9.7% (2.0-25.8) | 6.9% (2.3-15.5) | 9.4% (7.0-12.3) |
| Went to hospital | 2.8% (1.4-5.0) |  | 2.8% (0.3-9.7) | 2.7% (1.4-4.5) |
| Unknown | 2.3% (1.1-4.4) |  | 1.4% (0.0-7.5) | 2.0% (1.0-3.7) |
| **Reportable** |  |  |  |  |
| Yes | 2.6% (1.2-4.7) | 0% | 0% | 2.0% (1.0-3.7) |
| No | 97.4% (95.3-98.8) | 100% | 100% | 98.0% (96.3-99.0) |

### Large Animal Practice Employee Reported Incidents

There were 100 incidents recorded within SafetyHub, 87.0% (n=87) involved employees. Due to the low number of injuries sustained to non-employees, they were excluded from analysis. To achieve meaningful analysis we aggregated roles into three groups namely; Operations and Administration (n=19) (i.e. administrators, hospital directors, lab technicians, receptionists), Animal Care Roles (n=21) (i.e. veterinary nurses, animal care assistants, groomers), and veterinary surgeons (n=47). Over half of injuries reported were by veterinarians. We could only check representation of the veterinarians. They were broadly representative with 69.8% being female (66.8% nationally) and were a younger population with 37.2% being less than 30 years old (25.1% nationally) Table S6 (Surgeons, 2025).

Incidents occurred in a bimodal distribution, with peaks occurring between 9-11am and at 3-5pm (Fig1). Injuries primarily occurred in the consulting room (14.9%), in a stable or yard (13.8%), and on a farm (9.2%). (Table 7) This did vary by role, for example Operations and Administration staff were injured more in the lab (15.8%). However, there were a high number of reports where the location could not be classified or remained unknown (36.8%). Injuries predominately involved animals (73.6%), with horses being involved in over half of all injuries.

**Table 7.** Context of reported injuries to employees in UK large animal veterinary practices.

|  | Operations and Administration (n=19) | Animal Care Roles (n=21) | Veterinary Surgeons (n=47) | All staff roles (n=87) |
| --- | --- | --- | --- | --- |
| **Location of Injury** |  |  |  |  |
| Consult | 5.3% (0.1-26.0) | 19.0% (5.4-41.9) | 17.0% (7.6-30.8) | 14.9% (8.2-24.2) |
| Stables/Yard | 5.3% (0.1-26.0) | 28.6% (11.3-52.2) | 10.6% (3.5-23.1) | 13.8% (7.3-22.9) |
| Farm | 5.3% (0.1-26.0) |  | 14.9% (6.2-28.3) | 9.2% (4.1-17.3) |
| Car park | 5.3% (0.1-26.0) | 4.8% (0.1-23.8) | 4.3% (0.5-14.5) | 4.6% (1.3-11.4) |
| Prep | 5.3% (0.1-26.0) | 4.8% (0.1-23.8) | 4.3% (0.5-14.5) | 4.6% (1.3-11.4) |
| Imaging |  | 14.3% (3.0-36.3) | 2.1% (0.1-11.3) | 4.6% (1.3-11.4) |
| Lab | 15.8% (3.4-39.6) |  |  | 3.4% (0.7-9.7) |
| Office | 10.5% (1.3-33.1) |  |  | 2.3% (0.3-8.1) |
| Theatre | 5.3% (0.1-26.0) | 4.8% (0.1-23.8) |  | 2.3% (0.3-8.1) |
| Reception | 5.3% (0.1-26.0) |  | 2.1% (0.1-11.3) | 2.3% (0.3-8.1) |
| Lunging pen |  | 4.8% (0.1-23.8) |  | 1.1% (0.0-6.2) |
| Other | 31.6% (12.6-56.6) | 14.3% (3.0-36.3) | 27.7% (15.6-42.6) | 25.3% (16.6-35.8) |
| Unknown | 5.3% (0.1-26.0) | 4.8% (0.1-23.8) | 17.0% (7.6-30.8) | 11.5% (5.7-20.1) |
| **Animal involvement in injury** | **31.6% (12.6-56.6)** | **81.0% (58.1-94.6)** | **87.2% (74.3-95.2)** | **73.6% (63.0-82.5)** |
| Horse | 21.1% (6.1-45.6) | 71.4% (47.8-88.7) | 59.6% (44.3-73.6) | 54.0% (43.0-64.8) |
| Cow |  |  | 12.8% (4.8-25.7) | 6.9% (2.6-14.4) |
| Dog |  | 4.8% (0.1-23.8) | 8.5% (2.4-20.4) | 5.7% (1.9-12.9) |
| Cat |  | 4.8% (0.1-23.8) | 4.3% (0.5-14.5) | 3.4% (0.7-9.7) |
| Other animal | 10.5% (1.3-33.1) |  | 2.1% (0.1-11.3) | 3.4% (0.7-9.7) |
| **Mechanism of Injury** |  |  |  |  |
| **Animal Injured Person \*** | **31.6% (12.6-56.6)** | **81.0% (58.1-94.6)** | **83.0% (69.2-92.4)** | **71.3% (60.6-80.5)** |
| Clinical Examination | 10.5% (1.3-33.1) | 14.3% (3.0-36.3) | 27.7% (15.6-42.6) | 20.7% (12.8-30.7) |
| Clinical Procedure (with sedation) |  | 4.8% (0.1-23.8) | 14.9% (6.2-28.3) | 9.2% (4.1-17.3) |
| Clinical Procedure – no sedation) |  | 14.3% (3.0-36.3) | 8.5% (2.4-20.4) | 8.0% (3.3-15.9) |
| Ultrasound |  | 4.8% (0.1-23.8) | 10.6% (3.5-23.1) | 6.9% (2.6-14.4) |
| Entering/leaving stable or horsebox |  | 14.3% (3.0-36.3) | 2.1% (0.1-11.3) | 4.6% (1.3-11.4) |
| Blood sampling | 5.3% (0.1-26.0) |  | 2.1% (0.1-11.3) | 2.3% (0.3-8.1) |
| Injecting IV – Cannula placement |  |  | 4.3% (0.5-14.5) | 2.3% (0.3-8.1) |
| Radiography |  | 4.8% (0.1-23.8) | 2.1% (0.1-11.3) | 2.3% (0.3-8.1) |
| Admin oral meds |  | 4.8% (0.1-23.8) |  | 1.1% (0.0-6.2) |
| Anaesthesia/induction/recovery | 5.3% (0.1-26.0) |  |  | 1.1% (0.0-6.2) |
| General petting |  |  | 2.1% (0.1-11.3) | 1.1% (0.0-6.2) |
| Grooming |  | 4.8% (0.1-23.8) |  | 1.1% (0.0-6.2) |
| Handling |  | 4.8% (0.1-23.8) |  | 1.1% (0.0-6.2) |
| Unknown | 10.5% (1.3-33.1) | 9.5% (1.2-30.4) | 8.5% (2.4-20.4) | 9.2% (4.1-17.3) |
| **Non-animal injury** | **68.4% (43.5-87.4)** | **19.0% (5.4-41.9)** | **17.0% (7.6-30.8)** | **28.7% (19.5-39.4)** |
| Struck by equipment/furniture | 15.8% (3.4-39.6) | 4.8% (0.1-23.8) | 4.3% (0.5-14.5) | 6.9% (2.6-14.4) |
| Cut by non-surgical sharp | 15.8% (3.4-39.6) |  | 2.1% (0.1-11.3) | 4.6% (1.3-11.4) |
| Cut by surgical sharp | 10.5% (1.3-33.1) |  | 2.1% (0.1-11.3) | 3.4% (0.7-9.7) |
| Manual handling | 5.3% (0.1-26.0) | 4.8% (0.1-23.8) | 2.1% (0.1-11.3) | 3.4% (0.7-9.7) |
| Struck by falling/moving object | 10.5% (1.3-33.1) |  | 2.1% (0.1-11.3) | 3.4% (0.7-9.7) |
| Contact with harmful substance | 5.3% (0.1-26.0) | 4.8% (0.1-23.8) |  | 2.3% (0.3-8.1) |
| Exposure to heat/steam | 5.3% (0.1-26.0) |  |  | 1.1% (0.0-6.2) |
| Needlestick |  |  | 2.1% (0.1-11.3) | 1.1% (0.0-6.2) |
| Slips/Trips |  | 4.8% (0.1-23.8) |  | 1.1% (0.0-6.2) |
| Vehicular accident |  |  | 2.1% (0.1-11.3) | 1.1% (0.0-6.2) |

The most prevalent mechanisms of injury were a clinical examination (20.7%), clinical procedure (17.2%), ultrasound examination (6.9%), and being struck by equipment/furniture (6.4%). There was not enough contextual information of these events to perform further descriptive analysis.

The most prevalent resultant injuries were a bruise to a leg (14.1%) and a cut to a hand (11.8%) (Table S7). Head injuries made up 17.9% of injuries, whilst there were 10.6% fractures, 1.2% lost consciousness, and 1.2% received a concussion. Around a quarter of individuals (25.6%) did not receive medical treatment for their injuries (Table 8). Most received first aid (55.8%), and 17.5% attended a hospital. The majority (97.7%) took no time off work for their injuries, and less than 10.5% were RIDDOR reportable.

**Table 8.** Consequences of workplace injuries to employees at large animal veterinary practices.

|  | Operations and Administration (n=19) | Animal Care Roles (n=20) | Veterinary Surgeons (n=47) | Total - 86 |
| --- | --- | --- | --- | --- |
| **Medical Treatment** |  |  |  |  |
| None | 21.1% (6.1-45.6) | 25.0% (8.7-49.1) | 27.7% (15.6-42.6) | 25.6% (16.8-36.1) |
| First aid | 73.7% (48.8-90.9) | 60.0% (36.1-80.9) | 46.8% (32.1-61.9) | 55.8% (44.7-66.5) |
| Primary Care |  | 5.0% (0.1-24.9) |  | 1.2% (0.0-6.3) |
| Minor Injury Unit |  | 5.0% (0.1-24.9) | 4.3% (0.5-14.5) | 3.5% (0.7-9.9) |
| Emergency Department | 5.3% (0.1-26.0) | 5.0% (0.1-24.9) | 21.3% (10.7-35.7) | 14.0% (7.4-23.1) |
| **Time off work** |  |  |  |  |
| None | 100% | 100% | 95.7% (85.5-99.5) | 97.7% (91.9-99.7) |
| <7 days |  |  |  |  |
| 7-14 days |  |  | 4.3% (0.5-14.5) | 2.3% (0.3-8.2) |
| 15-28 days |  |  |  |  |
| >28 days |  |  |  |  |
| **Reportable** |  |  |  |  |
| Yes |  | 10.0% (1.2-31.7) | 14.9% (6.2-28.3) | 10.5% (4.9-18.9) |
| No | 100% | 90.0% (68.3-98.8) | 85.1% (71.7-93.8) | 89.5% (81.1-95.1) |

## Discussion

This study highlights high levels of occupational injury under-reporting within the UK veterinary sector; for every 100 injuries 69 remained unreported to their employer. A recent systematic review explored individual reporting to management across multiple professions, and in twelve studies they found a weighted mean of under-reporting of 39% (range: 20-74%) (Kyung et al., 2023). When this was stratified by health care workers (including medical doctors and nurses) the mean was 38%, and considering solely needlesticks it was 31% (Kyung et al., 2023). No previous work has estimated under-reporting in the veterinary profession. Our results suggest that under-reporting in the veterinary profession is almost twice the level found in the medical profession.

Those employed in companion animal practices having significantly higher rates of under-reporting compared to those in large animal practices. In companion animal practices these rates are high across all job roles, except administrators whose estimates must be treated with caution due to their wide confidence intervals. Individuals within this study population have already identified that they did not report their injuries because reporting was ‘too much effort’, they were ‘too busy’ to fill in the report, and that the injury was an inevitable everyday hazard that was not worthy of reporting (Furtado et al., 2024; Tulloch et al., 2025b). In a minority of cases some forgot, or did not want to be perceived differently be colleagues. This partially mirrors research in healthcare workers, where under-reporting has been linked to time and effort required, lack of awareness about how to report, the perception that injuries are either minor or just part of the job, and fear of discrimination after reporting an injury (Kyung et al., 2023). In large animal practices under-reporting rates varied across job role. However, due to small sample sizes in the survey data the resultant estimates’ confidence estimates were wide. The only job roles with a degree of precision were production animal vets (64.4%) and equine veterinarians (40.0%). Due to the relative lack of precision compared to the companion animal practices it is challenging to know how truly different they are. Reasons for not reporting are similar to companion animal practices in that the injury was too minor to report, they did not know they needed to report, an injury is an everyday hazard of the job, they were too busy, and that reporting would make no difference (Furtado et al., 2024; Tulloch et al., 2025a).

There are other factors currently unexplored in the veterinary sector that could influence injury reporting behaviours. Individuals with more severe injuries are more likely to report an injury, however this does not appear to be the case in the companion animal veterinary sector (Orellana et al., 2021). This could be attributed to the cumulative number of work-related injuries sustained, as prior research indicates that individuals who experience multiple injuries are progressively less likely to report them (Orellana et al., 2021). Notably, over 25% of companion animal veterinarians and veterinary nurses report having sustained more than ten occupational injuries over the course of their careers (Tulloch et al., 2025b). Demographics are unlikely to have played a role as age, gender, and education show mixed and inconsistent effects on under-reporting. Knowledge of the reporting process can also increase the likelihood of an individual reporting an injury (Kyung et al., 2023). However, we are unaware of the knowledge of CVS’s employees regarding their reporting guidelines, training received, and their specific barriers perceived to injury reporting.

Improving individual injury reporting in the veterinary workplace requires addressing systematic barriers and individual behaviours, however further research to understand these barriers in the veterinary profession is needed. Research from other industries indicates that the reporting process must be easy and accessible both in practice and remotely, online usability is key for reporting to be successful (Bernard et al., 2022). Modalities could include mobile apps or QR codes on posters around the workplace. Survey results suggest that the veterinary industry may perceive themselves as too busy to fill in a report (Tulloch et al., 2025a, b), so quickly fillable forms with minimum fields should be prioritised. The reporting process needs to stress confidentiality, especially with particularly sensitive incidents (Nicholson and Tait, 2002). Regular employee training on what constitutes a reportable injury, why reporting matters, and the process of reporting, can help improve reporting rates and reduces injury rates (Burke et al., 2006). This training has been shown to be more impactful at improving reporting rates when real-world case examples are used, to make the risks relatable to each specific job role (Burke et al., 2006). Although educational training has an important role, it should be complemented by other strategies, as evidence of its standalone impact is limited (Dyreborg et al., 2022). Cultural and psychological barriers impact injury reporting rates in other industries therefore these barriers need to be identified and addressed, such as fostering a culture of ‘no blame’ so that staff won’t be punished or judged for reporting (Probst and Estrada, 2010). The importance of reporting minor injuries needs to be stressed as they can indicate wider safety issues. Reframe reporting as a patient and team safety tool, shifting from a ‘quiet fix’ to ‘noisy fixes’ that highlights safety incidents and their solutions to all colleagues (Probst and Estrada, 2010). Veterinary employees will prioritise looking after a patient or colleague over themselves (Irwin et al., 2019). Messaging could reflect this altruism by promoting that reporting will aid in keeping their colleagues, clients and patients safe. Visible action based on incident reports to show that reporting leads to change has demonstrated improvement in injury reporting (Biswas et al., 2022; Dyreborg et al., 2022); this could be either through policy changes or new equipment (i.e. hard hat policies, provision of new sharps bins). Staff should be regularly updated on reported injuries and how managers responded to them. These improvements should be celebrated (Probst and Estrada, 2010). Senior leaders must model the desired behaviour and must report their own injuries and discuss them openly to practice members (Biswas et al., 2022). They need to create an environment of shared accountability as well as individual responsibility. Team based incentives can also be encouraged to promote strong safety reporting cultures (Biswas et al., 2022). Injury reporting should be seen as part of continuous improvement and quality improvement (Gibson et al., 2025); as such annual injury audits should be used to identify gaps

### Companion Animal Practice Incidents

Injuries in companion animal practices occurred throughout the day with peaks in the morning and afternoon reflecting the busiest time of day for clinical consultations and procedures. Very few injuries were recorded happening during ‘out of hours’, this could be reflective of few injuries are occurring or that few are reported, but without further exploration it is unclear. Injuries primarily occur to veterinarians and veterinary nurses, within the prep room, consultation room, and kennels, and with cats and dogs being the main cause of injury. This is reflective of self-reported injuries (Tulloch et al., 2025b). The injuries caused by cats were similar to self-reported injuries, whereas those caused by dogs differed. For dog-related injuries there were less fractures and head injuries in the audit than reported in the survey. This is surprising as usually more severe injuries have higher rates of injury reporting. This could be indicative of the poor injury culture within veterinary practices that even severe injuries are not reported. Contrastingly, 1% of all injuries were a ‘specified injury’ that would be reportable to HSE, this is similar to the survey results, suggestive that the overall prevalence of serious injuries may have captured in the audit.

This study provides a comprehensive stratification of dog-related occupational injuries by breed. There were fewer pure-bred dogs (73.3%) in the audit than estimates of the national dog population (85.8%) (McMillan et al., 2024). The top breeds involved in injuries were broadly reflective of the national demographics, though there were relatively more incidents involving Staffordshire Bull Terriers (11% of incidents vs 5% of national population), Border Collies (10% vs 2% of national population). There were fewer incidents involving Labrador Retrievers (5% vs 10% of national population) and Cocker Spaniels (4% vs 7% of national population). Whether these breeds inherently pose more or less risk to veterinary staff than other breeds is yet to be seen. It is the authors opinion that the prevalence of breeds is solely reflective of level of exposure faced by veterinary staff.

The context of events preceding injuries differed between the audit and the self-reported survey. In both, clinical examinations were the most common event. However, the audit identified needlestick injuries, drug administration, and exposure to harmful substances as subsequent causes, whereas the survey highlighted animal restraint, induction or recovery from general anaesthesia, and needlesticks as the next most frequent contexts. A higher number of injuries ended up in attending a hospital or minor injury unit in the survey (19.4% veterinarians, 16.3% veterinary nurses than in the audit (6%). Differences could be explained by the variation in what people perceive as reportable or memorable and how SafetyHub is used in practice. The audit may have captured incidents that are procedurally emphasised, in particular those with regulatory implications or that involve hazardous substances. The surveys rely on personal recollection and interpretation of risk and so may describe events that made a lasting impression, even if they didn’t report them. There could also be normalisation of certain risks (such as restraining a fractious animal) (Tulloch et al., 2025a, b), and so they are unlikely to report them. These discrepancies show that the audit may not accurately reflect what is occurring in veterinary practice. This highlights the importance of improving reporting culture and clarifying what should be reported.

### Needlesticks

Around one in ten reported injuries were caused by needlesticks, similar to the overall reporting in the survey paper but lower than the one in six for self-reported by veterinarians (Tulloch et al., 2025b). There was a lower rate of injuries caused by poor needle-handling practice (19.2%) compared to the survey (32.0%). The active ingredients mentioned were consistent with both datasets, likely reflective of prevalence of usage within clinical practice. The lower reporting rates of needlestick injuries in veterinarians and the higher rates of poor handling practices in the survey suggest that a significant number of incidents go undocumented through formal channels, potentially due to normalisation of risk, time constraints, or a perception that such injuries are an expected part of the job (Tulloch et al., 2025b), or that they don’t even fulfil the definition of a workplace injury (Furtado et al., 2024). These finding are supported by prior research (Weese and Jack, 2008; Johnson and Fritschi, 2024). This under-reporting may mask systemic issues around unsafe practices, inadequate training, or insufficient safety protocols. Further research is needed due to apparent under-reporting of needlestick injuries, inconsistent risk awareness and safety practices, frequent exposure to high-risk pharmaceuticals, and a clear need for targeted, evidence-based interventions.

### Harmful substances

The data reveal concerning patterns in exposure to harmful substances within veterinary workplaces. These hazards have been previously identified, but their frequency has been less well described (Jeyaretnam and Jones, 2000; Reijula et al., 2003; Epp and Waldner, 2012b, a; Fowler et al., 2016; Scheftel et al., 2017). The prominence of pepper spray may reflect its inappropriate or poorly managed use in police handling animals, raising questions about the availability, training, and situational justification for its deployment. If an animal, that may have been exposed to pepper spray, is seen in a practice it is critical that all who handle it most wear appropriate personal protective equipment (PPE) (gloves and eye protection) to minimise the risk of irritation to their eyes. Sedative-related exposures are likely to arise from the handling or administration of pharmaceuticals, suggesting the need for stricter adherence to handling procedures, secure storage, and clearer protocols around PPE. Disinfectant-related incidents suggest that even standard cleaning practices pose meaningful risks when PPE, ventilation, or safe dilution procedures are not adequately enforced. These findings collectively emphasise the need for comprehensive chemical safety training, improved risk assessment, and greater awareness of the risks associated with exposure to harmful substances in veterinary settings.

### Companion Animal Practice Client Incidents

To our knowledge this is the first research to describe injuries occurring to clients attending companion animal veterinary practices. These injuries made up over 40% of CVS UK Ltd’s accident records. Many incidents were in clinical environments, yet they were poorly documented. The potential for litigation from client injuries should incentivise businesses to improve the recording and reporting of such incidents, ensuring transparency and demonstrating a proactive approach to safety and accountability (Oliver et al., 2008). Many of the falls, slips and trips were linked to modifiable environmental hazards, such as scales, uneven surfaces and steps. It is likely that simple targeted interventions, such as redesigning clinical environments, better positioning of scales, or embedding scales in the floor, could reduce the risks of these hazards. Medical episodes were rare but did include serious life-threatening events. This highlights the need for veterinary clinics to be able to provide basic life support and clear protocols for managing acute health emergencies on site.

### Large Animal Practice Incidents

There were relatively few incidents reported to SafetyHub, the majority were horse-related injuries to vets and as such this is where we will focus the discussion. The main events preceding injuries were equivalent to previous studies, namely clinical examinations and procedures (Fritschi et al., 2006; Parkin et al., 2018; Tulloch et al., 2025a). Injury patterns had a similar prevalence of head injuries, and fractures to self-reported rates, whilst levels of medical attention and hospital attendance were equivalent (Tulloch et al., 2025a). RIDDOR reportable injuries were comparable. Differences lay in time taken off work, with less than 5% taking time off work in the audit versus 18% in the survey. It appears that what is recorded in the audit is broadly representative of the context and consequences of injuries in equine veterinary practices. Differences in time off work may be either reflective of under-reporting of injuries to the official reporting system, or the survey not capturing minor injuries that may not have resulted in time off work. Care must be taken with interpreting these results, as study numbers are small and so may lack precision.

### Limitations

The major limitations of this study are that it was not possible to directly link injuries in the survey to records in the safety audit, and that the prevalence estimates from the survey assume that each respondent experienced only one injury per year. For the former point we could have attempted to match individuals on demographic characteristics and the contextual details of the injuries described in the survey. However, we deliberately chose not to pursue this approach, as we had committed to maintaining full anonymity for survey participants to encourage open and honest reporting. Seeking ethical approval for identifiable linkage would have compromised that assurance and, in our view, potentially hindered recruitment and reduced response rates. The latter point, assuming one injury per person per year, was necessary to generate conservative prevalence estimates. However, literature suggest that many veterinary professionals may experience multiple work-related injuries in a year (Johnson and Fritschi, 2024; Tulloch et al., 2025a, b). Therefore, our under-reporting estimate is likely to under-estimate the true level of under-reporting across the veterinary sector.

There was a demographic mismatch between those in the audit and the UK veterinary sector. It is unclear whether this reflects differences in the composition of the CVS workforce or whether certain groups are more likely to report injuries. There were more female and young reporters of accidents. As discussed above, in other industries it is unclear whether demographics have a part to be in reporting habits. These discrepancies raise important questions about the representativeness of accident book data and highlight the need for further investigation into reporting behaviours.

## Conclusions

This study highlights significant under-reporting of occupational injuries in the UK veterinary sector, particularly within companion animal practices, where nearly 70% of injuries go unreported. Barriers include time constraints, normalisation of injury, and unclear reporting processes. Addressing this requires streamlined digital reporting, clear training, leadership modelling, and a visible response to reported incidents. Enhancing safety culture and injury surveillance is essential to protect veterinary professionals and ensure workplace risks are adequately addressed.

## Data Availability

All data produced in the present study are available upon reasonable request to the authors

## Declaration of generative AI and AI-assisted technologies in the writing process

During the preparation of this work the authors used ChatGPT-4 to improve the language and readability of the manuscript. After using this tool, the authors reviewed and edited the content as needed and take full responsibility for the content of the published article.

## Author Contributions

Conceptualisation: JT. Funding Acquisition – JT, MW, RJ. Methodology: JT, MW, IS. Formal analysis: JT. Writing original draft – JT. Writing review & editing – All authors

## References

Bernard, R.M., Toppo, C., Raggi, A., de Mul, M., de Miquel, C., Pugliese, M.T., van der Feltz-Cornelis, C.M., Ortiz-Tallo, A., Salvador-Carulla, L., Lukersmith, S., Hakkaart-van Roijen, L., Merecz-Kot, D., Staszewska, K., Sabariego, C., 2022. Strategies for Implementing Occupational eMental Health Interventions: Scoping Review. J Med Internet Res 24, e34479.

Biswas, A., Begum, M., Van Eerd, D., Johnston, H., Smith, P.M., Gignac, M.A.M., 2022. Integrating Safety and Health Promotion in Workplaces: A Scoping Review of Facilitators, Barriers, and Recommendations. Health Promot Pract 23, 984–998.

Boden, L.I., Ozonoff, A., 2008. Capture-recapture estimates of nonfatal workplace injuries and illnesses. Ann Epidemiol 18, 500–506.

Burke, M.J., Sarpy, S.A., Smith-Crowe, K., Chan-Serafin, S., Salvador, R.O., Islam, G., 2006. Relative effectiveness of worker safety and health training methods. Am J Public Health 96, 315–324.

Dyreborg, J., Lipscomb, H.J., Nielsen, K., Torner, M., Rasmussen, K., Frydendall, K.B., Bay, H., Gensby, U., Bengtsen, E., Guldenmund, F., Kines, P., 2022. Safety interventions for the prevention of accidents at work: A systematic review. Campbell Syst Rev 18, e1234.

Epp, T., Waldner, C., 2012a. Occupational health hazards in veterinary medicine: physical, psychological, and chemical hazards. Can Vet J 53, 151–157.

Epp, T., Waldner, C., 2012b. Occupational health hazards in veterinary medicine: zoonoses and other biological hazards. Can Vet J 53, 144–150.

Executive, H.a.S., 2013. Reporting of Injuries, Diseases and Dangerous Occurrences Regulations 2013 - RIDDOR - HSE.

Fowler, H.N., Holzbauer, S.M., Smith, K.E., Scheftel, J.M., 2016. Survey of occupational hazards in Minnesota veterinary practices in 2012. J Am Vet Med Assoc 248, 207–218.

Fritschi, L., Day, L., Shirangi, A., Robertson, I., Lucas, M., Vizard, A., 2006. Injury in Australian veterinarians. Occup Med (Lond) 56, 199–203.

Furtado, T., Whiting, M., Schofield, I., Jackson, R., Tulloch, J.S.P., 2024. Pain, inconvenience and blame: defining work-related injuries in the veterinary workplace. Occup Med (Lond) 74, 501–507.

Gabel, C.L.,, Gerberich, S.G.,, 2002. Risk factors for injury among veterinarians. 13, 80–86.

Gibson, J., Reyneke, R.A., Basham, N., Rooke, F., Morrow, L., Richens, I.F., Messina, D., Brennan, M.L., 2025. Evidence-based veterinary medicine and quality improvement: ‘peas in a pod’ for improving patient care. Vet Rec 196, 267–270.

Irwin, A., Vikman, J., Ellis, H., 2019. ‘No-one knows where you are’: veterinary perceptions regarding safety and risk when alone and on-call. Vet Rec 185, 728.

Jeyaretnam, J., Jones, H., 2000. Physical, chemical and biological hazards in veterinary practice. Aust Vet J 78, 751–758.

Johnson, L., Fritschi, L., 2024. Frequency of workplace incidents and injuries in veterinarians, veterinary nurses and veterinary students and measures to control these. Aust Vet J 102, 431–439.

Kyung, M., Lee, S.J., Dancu, C., Hong, O., 2023. Underreporting of workers’ injuries or illnesses and contributing factors: a systematic review. BMC Public Health 23, 558.

Lipscomb, H.J., Schoenfisch, A.L., Cameron, W., 2015. Non-reporting of work injuries and aspects of jobsite safety climate and behavioral-based safety elements among carpenters in Washington State. Am J Ind Med 58, 411–421.

McMillan, K.M., Harrison, X.A., Wong, D.C., Upjohn, M.M., Christley, R.M., Casey, R.A., 2024. Estimation of the size, density, and demographic distribution of the UK pet dog population in 2019. Sci Rep 14, 31746.

Nicholson, A.N., Tait, P.C., 2002. Confidential reporting: from aviation to clinical medicine. Clin Med (Lond) 2, 234–236.

Nienhaus, A., Skudlik, C., Seidler, A., 2005. Work-related accidents and occupational diseases in veterinarians and their staff. Int Arch Occup Environ Health 78, 230–238.

Oliver, D., Killick, S., Even, T., Willmott, M., 2008. Do falls and falls-injuries in hospital indicate negligent care -- and how big is the risk? A retrospective analysis of the NHS Litigation Authority Database of clinical negligence claims, resulting from falls in hospitals in England 1995 to 2006. Qual Saf Health Care 17, 431–436.

Orellana, C., Kreshpaj, B., Burstrom, B., Davis, L., Frumento, P., Hemmingsson, T., Johansson, G., Kjellberg, K., Wegman, D.H., Bodin, T., 2021. Organisational factors and under-reporting of occupational injuries in Sweden: a population-based study using capture-recapture methodology. Occup Environ Med 78, 745–752.

Parkin, T.D.H., Brown, J., Macdonald, E.B., 2018. Occupational risks of working with horses: A questionnaire survey of equine veterinary surgeons. Equine Veterinary Education 30, 200–205.

plc, C.G., 2024. CVS Group plc Annual Report and Financial Statements for the year ended 30 June 2024.

Probst, T.M., Brubaker, T.L., Barsotti, A., 2008. Organizational injury rate underreporting: the moderating effect of organizational safety climate. J Appl Psychol 93, 1147–1154.

Probst, T.M., Estrada, A.X., 2010. Accident under-reporting among employees: testing the moderating influence of psychological safety climate and supervisor enforcement of safety practices. Accid Anal Prev 42, 1438–1444.

Reijula, K., Rasanen, K., Hamalainen, M., Juntunen, K., Lindbohm, M.L., Taskinen, H., Bergbom, B., Rinta-Jouppi, M., 2003. Work environment and occupational health of Finnish veterinarians. Am J Ind Med 44, 46–57.

Riley, C.B., Liddiard, J.R., Thompson, K., 2015. A Cross-Sectional Study of Horse-Related Injuries in Veterinary and Animal Science Students at an Australian University. Animals (Basel) 5, 951–964.

Rosenman, K.D., Kalush, A., Reilly, M.J., Gardiner, J.C., Reeves, M., Luo, Z., 2006. How much work-related injury and illness is missed by the current national surveillance system? J Occup Environ Med 48, 357–365.

Scheftel, J.M., Elchos, B.L., Rubin, C.S., Decker, J.A., 2017. Review of hazards to female reproductive health in veterinary practice. J Am Vet Med Assoc 250, 862–872.

Surgeons, R.C.o.V., 2025. RCVS Facts 2024.

Tulloch, J.S.P., Fleming, K.M., Pinchbeck, G., Forster, J., Lowe, W., Westgarth, C., 2023. Audit of animal-related injuries at UK veterinary schools between 2009 and 2018. Vet Rec 193, e3171.

Tulloch, J.S.P., Schofield, I., Jackson, R., Whiting, M., 2025a. ‘It’s only a flesh wound’ - Understanding the safety culture in equine, production animal and mixed veterinary practices. Prev Vet Med 241, 106541.

Tulloch, J.S.P., Schofield, I., Jackson, R., Whiting, M., 2025b. ‘Just part of the job’ - understanding work-related injuries and safety culture in companion animal veterinary practices. J Small Anim Pract. 10.1111/jsap.70039

Voss, D.S., Boyd, M.V., Evanson, J.F., Bender, J.B., 2024. An increase in animal-related occupational injuries at a veterinary medical center (2008-2022). J Am Vet Med Assoc 262, 376–382.

Weese, J.S., Jack, D.C., 2008. Needlestick injuries in veterinary medicine. Can Vet J 49, 780–784.

