## Supplementary Material for "Hidden Harm: Quantifying occupational injury under-reporting in veterinary workplaces through modified capture-recapture analysis"

### Companion Animal Practice Incidents

**Table S1** – Demographics of companion animal staff reporting a work-related injury

| | Administrative staff (n=34) (95% CI) | Reception Staff (n=44) (95% CI) | Animal Care Assistants (n=89) (95% CI) | Student Veterinary Nurses (n=83) (95% CI) | Veterinary Nurses (n=243) (95% CI) | Veterinary Surgeons (n=152) (95% CI) |
|---|---|---|---|---|---|---|
| **Gender (% Female)** | N=32 87.5% (71.0-96.5) | N=42 100% | N=74 100% | N=75 98.7% (92.8-99.9) | N=221 95.9% (92.4-98.1) | N=134 79.1% (71.2-85.6) |
| **Age** | N=32 | N=42 | N=74 | N=75 | N=219 | N=132 |
| **<20** | | | 4.1% (0.8-11.4) | | | |
| **20-29** | 15.6% (5.3-32.8) | 16.7% (7.0-31.4) | 48.6% (36.9-60.6) | 64.0% (52.1-74.8) | 38.4% (31.9-45.2) | 47.0% (38.2-55.9) |
| **30-39** | 28.1% (13.8-46.8) | 28.6% (15.7-44.6) | 28.4% (18.5-40.1) | 33.3% (22.9-45.2) | 41.1% (34.5-47.9) | 26.5% (19.2-34.9) |
| **40-49** | 31.3% (16.1-50.0) | 19.0% (8.6-34.1) | 9.5% (3.9-18.5) | 2.7% (0.3-9.3) | 16.0% (11.4-21.5) | 14.4% (8.9-21.6) |
| **50-59** | 25.0% (11.5-43.3) | 23.8% (12.1-39.5) | 6.8% (2.2-15.1) | | 3.7% (1.6-7.1) | 9.8% (5.3-16.3) |
| **60+** | | 11.9% (4.0-25.6) | 2.7% (0.3-9.4) | | 0.9% (0.1-3.3) | 2.3% (0.5-6.5) |

**Table S2** – Types of harmful substances reported in companion animal accident books

| Substance | Employees (n=41) 95% CI |
|---|---|
| Pepper Spray | 24.4% (12.4-40.3) |
| Pentobarbital | 14.6% (5.6-29.2) |
| G9 Disinfectant (mix of QAC* and Chlorohexidine) | 12.2% (4.1-26.2) |
| Lidocaine | 7.3% (1.5-19.9) |
| Kennel cough vaccine | 4.9% (0.6-16.5) |
| Premed (unspecified) | 4.9% (0.6-16.5) |
| Sterilium (alcohol-based disinfectant) | 4.9% (0.6-16.5) |
| Bupivacaine Hydrochloride | 2.4% (0.1-12.9) |
| Diff-Quik solution A | 2.4% (0.1-12.9) |
| Exigo diluent | 2.4% (0.1-12.9) |
| Hand sanitizer | 2.4% (0.1-12.9) |
| Hibiscrub | 2.4% (0.1-12.9) |
| Ketamine | 2.4% (0.1-12.9) |
| Medetomidine/buprenorphine | 2.4% (0.1-12.9) |
| Methadone | 2.4% (0.1-12.9) |
| Metoclopramide | 2.4% (0.1-12.9) |
| Propofol | 2.4% (0.1-12.9) |
| Vaccine (unspecified) | 2.4% (0.1-12.9) |

*Quaternary ammonium chloride

**Table S3 –** Types of reported injuries and affected anatomical sites to employees in companion animal practices

| | | Cat | Dog | Other Animal | Non-animal-related | Total |
|---|---|---|---|---|---|---|
| | | n=217 | n=160 | n=23 | n=231 | N=631 |
| **Puncture wound** | | **50.2% (43.4-57.1)** | **38.8% (31.2-46.8)** | **65.2% (42.7-83.6)** | **25.1% (19.7-31.2)** | **38.7% (34.9-42.6)** |
| | Arm | 4.6% (2.2-8.3) | 3.1% (1.0-7.2) | | 1.3% (0.3-3.7) | 2.9% (1.7-4.5) |
| | Face | 0.5% (0.0-2.5) | 1.3% (0.2-4.4) | | 0.9% (0.1-3.1) | 0.8% (0.3-1.8) |
| | Hand | 43.8% (37.1-50.7) | 32.5% (25.3-40.4) | 60.9% (38.5-80.3) | 21.2% (16.1-27.1) | 33.3% (29.6-37.1) |
| | Leg | | 1.9% (0.4-5.4) | | 1.7% (0.5-4.4) | 1.1% (0.4-2.3) |
| | Wrist | 1.4% (0.3-4.0) | | 4.3% (0.1-22.0) | | 0.6% (0.2-1.6) |
| **Scratch** | | **39.2% (32.6-46.0)** | **22.5% (16.3-29.8)** | **17.4% (5.0-38.8)** | **9.5% (6.1-14.1)** | **23.3% (20.1-26.8)** |
| | Ankle | 0.5% (0.0-2.5) | | | | 0.2% (0.0-0.9) |
| | Arm | 6.0% (3.2-10.0) | 8.1% (4.4-13.5) | 8.7% (1.1-28.0) | 1.7% (0.5-4.4) | 5.1% (3.5-7.1) |
| | Back | | 0.6% (0.0-3.4) | | | 0.2% (0.0-0.9) |
| | Face | 3.2% (1.3-6.5) | 1.9% (0.4-5.4) | | | 1.6% (0.8-2.9) |
| | Hand | 27.2% (21.4-33.6) | 7.5% (3.9-12.7) | 8.7% (1.1-28.0 | 6.1% (3.4-10.0) | 13.8% (11.2-16.7) |
| | Head | | | | 0.9% (0.1-3.1) | 0.3% (0.0-1.1) |
| | Leg | | 1.3% (0.2-4.4) | | | 0.3% (0.0-1.1) |
| | Neck | 0.5% (0.0-2.5) | 0.6% (0.0-3.4) | | | 0.3% (0.0-1.1) |
| | Torso | | 0.6% (0.0-3.4) | | | 0.2% (0.0-0.9) |
| | Wrist | 1.8% (0.5-4.7) | 1.9% (0.4-5.4) | | 0.9% (0.1-3.1) | 1.4% (0.7-2.7) |
| **Bruise** | | **3.7% (1.6-7.1)** | **23.1% (16.8-30.4)** | **4.3% (0.1-22.0)** | **15.6% (11.2-20.9)** | **13.3% (10.8-16.2)** |
| | Ankle | | | | 0.4% (0.0-2.4) | 0.2% (0.0-0.9) |
| | Arm | | 6.3% (3.0-11.2) | | 0.4% (0.0-2.4) | 1.7% (0.9-3.1) |
| | Back | | 1.3% (0.2-4.4) | | 0.9% (0.1-3.1) | 0.6% (0.2-1.6) |
| | Eye | | | | 0.4% (0.0-2.4) | 0.2% (0.0-0.9) |
| | Face | | 0.6% (0.0-3.4) | | 2.6% (1.0-5.6) | 1.1% (0.4-2.3) |
| | Foot | | | | 1.3% (0.3-3.7) | 0.5% (0.1-1.4) |
| | Hand | 3.7% (1.6-7.1) | 10.6% (6.3-16.5) | 4.3% (0.1-22.0) | 3.5% (1.5-6.7) | 5.4% (3.8-7.4) |
| | Head | | 1.3% (0.2-4.4) | | 4.3% (2.1-7.8) | 1.9% (1.0-3.3) |
| | Hip | | 0.6% (0.0-3.4) | | 0.4% (0.0-2.4) | 0.3% (0.0-1.1) |
| | Leg | | 0.6% (0.0-3.4) | | 1.3% (0.3-3.7) | 0.6% (0.2-1.6) |
| | Neck | | 0.6% (0.0-3.4) | | | 0.2% (0.0-0.9) |
| | Torso | | 0.6% (0.0-3.4) | | | 0.2% (0.0-0.9) |
| | Wrist | | 0.6% (0.0-3.4) | | | 0.2% (0.0-0.9) |
| **Cut** | | **6.5% (3.6-10.6)** | **5.0% (2.2-9.6)** | **13.0% (2.8-33.6)** | **16.9% (12.3-22.4)** | **10.1% (7.9-12.8)** |
| | Arm | 0.5% (0.0-2.5) | 0.6% (0.0-3.4) | | 0.4% (0.0-2.4) | 0.5% (0.1-1.4) |
| | Eye | | | | 0.9% (0.1-3.1) | 0.3% (0.0-1.1) |
| | Hand | 6.0% (3.2-10.0) | 2.5% (0.7-6.3) | 13.0% (2.8-33.6) | 13.4% (9.3-18.5) | 8.1% (6.1-10.5) |
| | Head | | | | 2.2% (0.7-5.0) | 0.8% (0.3-1.8) |
| | Leg | | 0.6% (0.0-3.4) | | | 0.2% (0.0-0.9) |
| | Wrist | | 1.3% (0.2-4.4) | | | 0.3% (0.0-1.1) |
| **Chemical Irritation** | | | **0.6% (0.0-3.4)** | | **15.2% (10.8-20.4)** | **5.7% (4.0-7.8)** |
| | Eyes | | 0.6% (0.0-3.4) | | 10.4% (6.8-15.1) | 4.0% (2.6-5.8) |
| | Face | | | | 3.5% (1.5-6.7) | 1.3% (0.5-2.5) |
| | Hand | | | | 0.4% (0.0-2.4) | 0.2% (0.0-0.9) |
| | Torso | | | | 0.9% (0.1-3.1) | 0.3% (0.0-1.1) |
| **Musculoskeletal Injury (ie sprain)** | | | **6.9% (3.5-12.0)** | | **8.2% (5.0-12.6)** | **4.8% (3.2-6.7)** |
| | Ankle | | | | 1.7% (0.5-4.4) | 0.6% (0.2-1.6) |
| | Arm | | 0.6% (0.0-3.4) | | 1.7% (0.5-4.4) | 0.8% (0.3-1.8) |
| | Back | | 1.3% (0.2-4.4) | | 3.5% (1.5-6.7) | 1.6% (0.8-2.9) |
| | Hand | | 1.9% (0.4-5.4) | | 0.4% (0.0-2.4) | 0.6% (0.2-1.6) |
| | Neck | | 0.6% (0.0-3.4) | | | 0.2% (0.0-0.9) |
| | Shoulder | | 2.5% (0.7-6.3) | | | 0.6% (0.2-1.6) |
| | Wrist | | | | 0.9% (0.1-3.1) | 0.3% (0.0-1.1) |
| **Burn** | | | | | **3.9% (1.8-7.3)** | **1.4% (0.7-2.7)** |

| | | | | | | |
|---|---|---|---|---|---|---|
| | Arm | | | | 0.9% (0.1-3.1) | 0.3% (0.0-1.1) |
| | Hand | | | | 3.0% (1.2-6.1) | 1.1% (0.4-2.3) |
| **Fracture** | | | **1.9% (0.4-5.4)** | | **0.9% (0.1-3.1)** | **0.8% (0.3-1.8)** |
| | Fingers | | 0.6% (0.0-3.4) | | | 0.2% (0.0-0.9) |
| | Hand | | 1.3% (0.2-4.4) | | 0.4% (0.0-2.4) | 0.5% (0.1-1.4) |
| | Wrist | | | | 0.4% (0.0-2.4) | 0.2% (0.0-0.9) |
| **Contamination** | | **0.5% (0.0-2.5)** | **1.3% (0.2-4.4)** | | **0.4% (0.0-2.4)** | **0.6% (0.2-1.6)** |
| | Arm | | | | 0.4% (0.0-2.4) | 0.2% (0.0-0.9) |
| | Eyes | | 1.3% (0.2-4.4) | | | 0.3% (0.0-1.1) |
| | Face | 0.5% (0.0-2.5) | | | | 0.2% (0.0-0.9) |
| **Electric Shock** | | | | | **1.3% (0.3-3.7)** | **0.5% (0.1-1.4)** |
| | Arm | | | | 0.4% (0.0-2.4) | 0.2% (0.0-0.9) |
| | Hand | | | | 0.9% (0.1-3.1) | 0.3% (0.0-1.1) |
| **Crush** | | | | | **1.3% (0.3-3.7)** | **0.5% (0.1-1.4)** |
| | Back | | | | 0.4% (0.0-2.4) | 0.2% (0.0-0.9) |
| | Hand | | | | 0.9% (0.1-3.1) | 0.3% (0.0-1.1) |
| **Allergic Response** | | | | | **0.9% (0.1-3.1)** | **0.3% (0.0-1.1)** |
| | Face | | | | 0.4% (0.0-2.4) | 0.2% (0.0-0.9) |
| | Neck | | | | 0.4% (0.0-2.4) | 0.2% (0.0-0.9) |
| **Loss of consciousness** | | | | | **0.4% (0.0-2.4)** | **0.2% (0.0-0.9)** |
| **Radiation exposure** | | | | | **0.4% (0.0-2.4)** | **0.2% (0.0-0.9)** |
| | Hand | | | | 0.4% (0.0-2.4) | 0.2% (0.0-0.9) |

Table S4 – Dog breeds involved in dog-related injuries in companion animal veterinary practices

| **Breed** | n=135 |
|---|---|
| **Unknown cross-bred** | **15.6%** |
| **Other non-KC breeds** | **11.1%** |
| **Kennel Club Recognised Breeds** | **73.3%** |
| **Small-sized breeds** | **35.6%** |
| Staffordshire Bull Terrier | 11.1% |
| French Bulldog | 5.2% |
| Cocker Spaniel | 3.7% |
| Beagle | 2.2% |
| Bichon Frise | 2.2% |
| Dachshund | 1.5% |
| Terrier - Jack Russell | 1.5% |
| Shih-tzu | 1.5% |
| Terrier - West Highland W | 1.5% |
| Chihuahua | 0.7% |
| Lhasa Apso | 0.7% |
| Maltese | 0.7% |
| Pug | 0.7% |
| Spaniel - Cavalier King Charles | 0.7% |
| Terrier - Border (Border | 0.7% |
| Terrier - Yorkshire (York | 0.7% |
| **Large-sized breeds** | **28.1%** |
| Border Collie | 9.6% |
| Labrador Retriever | 5.2% |
| German Shepherd | 3.7% |
| Doberman | 1.5% |
| Greyhound | 1.5% |
| Maremma Sheepdog | 1.5% |
| Rottweiler | 1.5% |
| Akita | 0.7% |
| Boxer | 0.7% |
| Mastiff | 0.7% |
| Newfoundland | 0.7% |
| Retriever - Golden | 0.7% |
| **Medium-sized breeds** | **6.7%** |
| Springer Spaniel | 3.7% |
| Shar-Pei | 1.5% |
| Samoyed | 0.7% |
| Schnauzer | 0.7% |
| **Medium-large sized breeds** | **1.5%** |
| Husky | 1.5% |
| **Extra-large sized breeds** | **1.5%** |
| Saint Bernard | 0.7% |
| Wolfhound | 0.7% |

**Table S5 –** Types of reported injuries and affected anatomical sites to non-employees in companion animal practices

| | | Cat | Dog | Other Animal | Non-animal related | Total |
|---|---|---|---|---|---|---|
| | | **n=208** | **n=133** | **n=6** | **n=115** | **n=462** |
| Scratch | | **43.8% (36.9-50.8)** | **41.4% (32.9-50.2)** | **16.7% (0.4-64.1)** | **16.5% (10.3-24.6)** | **35.9% (31.6-40.5)** |
| | Hand | 34.1% (27.7-41.0) | 17.3% (11.3-24.8) | 16.7% (0.4-64.1) | 8.7% (4.3-15.4) | 22.7% (19.0-26.8) |
| | Arm | 4.3% (2.0-8.1) | 16.5% (10.7-24.0) | | 0.9% (0.0-4.8) | 6.9% (4.8-9.6) |
| | Face | 3.4% (1.4-6.8) | | | 0.9% (0.0-4.8) | 1.7% (0.8-3.4) |
| | Head | | 1.5% (0.2-5.3) | | 2.6% (0.5-7.4) | 1.1% (0.4-2.5) |
| | Wrist | 1.0% (0.1-3.4) | 2.3% (0.5-6.5) | | | 1.1% (0.4-2.5) |
| | Leg | | 0.8% (0.0-4.1) | | 1.7% (0.2-6.1) | 0.6% (0.1-1.9) |
| | Torso | 1.0% (0.1-3.4) | 0.8% (0.0-4.1) | | | 0.6% (0.1-1.9) |
| | Neck | | 1.5% (0.2-5.3) | | | 0.4% (0.1-1.6) |
| | Ankle | | | | 0.9% (0.0-4.8) | 0.2% (0.0-1.2) |
| | Back | | | | 0.9% (0.0-4.8) | 0.2% (0.0-1.2) |
| | Unknown | | 0.8% (0.0-4.1) | | | 0.2% (0.0-1.2) |
| Puncture wound | | **42.3% (35.5-49.3)** | **30.8% (23.1-39.4)** | **50.0% (11.8-88.2)** | **18.3% (11.7-26.6)** | **33.1% (28.8-37.6)** |
| | Hand | 38.5% (31.9-45.4) | 21.1% (14.5-29.0) | 16.7% (0.4-64.1) | 18.3% (11.7-26.6) | 28.1% (24.1-32.5) |
| | Arm | 0.5% (0.0-2.7) | 5.3% (2.1-10.5) | 33.3% (4.3-77.7) | | 2.2% (1.0-3.9) |
| | Wrist | 1.9% (0.5-4.9) | | | | 0.9% (0.2-2.2) |
| | leg | | 2.3% (0.5-6.5) | | | 0.6% (0.1-1.9) |
| | Face | | 1.5% (0.2-5.3) | | | 0.4% (0.1-1.6) |
| | Head | | 0.8% (0.0-4.1) | | | 0.2% (0.0-1.2) |
| | Hip | 0.5% (0.0-2.7) | | | | 0.2% (0.0-1.2) |
| | Unknown | 1.0% (0.1-3.4) | | | | 0.4% (0.1-1.6) |
| Cut | | **13.5% (9.1-18.9)** | **13.5% (8.2-20.5)** | **33.3% (4.3-77.7)** | **23.5% (16.1-32.3)** | **16.2% (13.0-19.9)** |
| | Hand | 13.0% (8.7-18.3) | 9.0% (4.7-15.2) | 33.3% (4.3-77.7) | 11.3% (6.2-18.6) | 11.7% (8.9-15.0) |
| | Face | 0.5% (0.0-2.7) | 3.8% (1.2-8.6) | | 9.6% (4.9-16.5) | 3.7% (2.2-5.8) |
| | Arm | | 0.8% (0.0-4.1) | | 0.9% (0.0-4.8) | 0.4% (0.1-1.6) |
| | Foot | | | | 0.9% (0.0-4.8) | 0.2% (0.0-1.2) |
| | Leg | | | | 0.9% (0.0-4.8) | 0.2% (0.0-1.2) |
| Bruise | | **0.5% (0.0-2.7)** | **10.5% (5.9-17.0)** | | **18.3% (11.7-26.6)** | **7.8% (5.5-10.6)** |
| | Head | | 2.3% (0.5-6.5) | | 6.1% (2.5-12.1) | 2.2% (1.0-3.9) |
| | Hand | 0.5% (0.0-2.7) | 4.5% (1.7-9.6) | | 1.7% (0.2-6.1) | 1.9% (0.9-3.7) |
| | Face | | 1.5% (0.2-5.3) | | 4.3% (1.4-9.9) | 1.5% (0.6-3.1) |
| | Arm | | 2.3% (0.5-6.5) | | 1.7% (0.2-6.1) | 1.1% (0.4-2.5) |
| | Foot | | | | 1.7% (0.2-6.1) | 0.4% (0.1-1.6) |
| | Leg | | | | 1.7% (0.2-6.1) | 0.4% (0.1-1.6) |
| | Ankle | | | | 0.9% (0.0-4.8) | 0.2% (0.0-1.2) |
| Loss of consciousness | | | | | **7.8% (3.6-14.3)** | **1.9% (0.9-3.7)** |
| Chemical irritation | | | | | **7.0% (3.1-13.3)** | **1.7% (0.8-3.4)** |
| | Eyes | | | | 7.0% (3.1-13.3) | **1.7% (0.8-3.4)** |
| Musculoskeletal Injury (ie sprain) | | | **1.5% (0.2-5.3)** | | **3.5% (1.0-8.7)** | **1.3% (0.5-2.8)** |
| | Wrist | | 0.8% (0.0-4.1) | | 1.7% (0.2-6.1) | 0.6% (0.1-1.9) |
| | Arm | | | | 0.9% (0.0-4.8) | 0.2% (0.0-1.2) |
| | Back | | | | 0.9% (0.0-4.8) | 0.2% (0.0-1.2) |
| | Shoulder | | 0.8% (0.0-4.1) | | | 0.2% (0.0-1.2) |
| Contamination | | | **1.5% (0.2-5.3)** | | **1.7% (0.2-6.1)** | **0.9% (0.2-2.2)** |
| | Face | | 1.5% (0.2-5.3) | | | 0.4% (0.1-1.6) |
| | Eyes | | | | 1.7% (0.2-6.1) | 0.4% (0.1-1.6) |
| Fracture | | | | | **2.6% (0.5-7.4)** | **0.6% (0.1-1.9)** |
| | Ankle | | | | 0.9% (0.0-4.8) | 0.2% (0.0-1.2) |
| | Arm | | | | 0.9% (0.0-4.8) | 0.2% (0.0-1.2) |
| | Hip | | | | 0.9% (0.0-4.8) | 0.2% (0.0-1.2) |
| Dislocation | | | **0.8% (0.0-4.1)** | | | **0.2% (0.0-1.2)** |
| | Shoulder | | 0.8% (0.0-4.1) | | | 0.2% (0.0-1.2) |
| Allergic Response | | | | | **0.9% (0.0-4.8)** | **0.2% (0.0-1.2)** |
| | Hand | | | | 0.9% (0.0-4.8) | 0.2% (0.0-1.2) |

### Large Animal Practice Employee Incidents

Table S6 Demographics of large animal staff reporting a work-related injury

| | **Operations and Administration (n=19)** | **Animal Care Roles (n=18)** | **Veterinary Surgeons (n=43)** | **All staff roles (n=80)** |
|---|---|---|---|---|
| **Sex** (Female) | 78.9% (54.4-94.0) | 100% | 69.8% (53.9-82.8) | 78.8% (68.2-87.1) |
| **Age** | | | | |
| 20-29 | 31.6% (12.6-56.6) | 72.2% (46.5-90.3) | 37.2% (23.0-53.3) | 43.8% (32.7-55.3) |
| 30-39 | 15.8% (3.4-39.6) | 16.7% (3.6-41.4) | 30.2% (17.2-46.1) | 23.8% (15.0-34.6) |
| 40-49 | 21.1% (6.1-45.6) | 5.6% (0.1-27.3) | 18.6% (8.4-33.4) | 16.3% (8.9-26.2) |
| 50-59 | 21.1% (6.1-45.6) | 5.6% (0.1-27.3) | 9.3% (2.6-22.1) | 11.3% (5.3-20.3) |
| 60+ | 10.5% (1.3-33.1) | | 4.7% (0.6-15.8) | 5.0% (1.4-12.3) |

**Table S7 –** Types of reported injuries and affected anatomical sites to employees in large animal practices

| | | **Horse (n=45)** | **Other Animals (n=17)** | **Non-animal related (n=23)** | **Total (n=85)** |
|---|---|---|---|---|---|
| Bruise | | **51.1% (35.8-66.3)** | **17.6% (3.8-43.4)** | **34.8% (16.4-57.3)** | **40.0% (29.5-51.2)** |
| | Leg | 24.4% (12.9-39.5) | 5.9% (0.1-28.7) | | 14.1% (7.5-23.4) |
| | Arm | 4.4% (0.5-15.2) | | 8.7% (1.1-28.0) | 4.7% (1.3-11.6) |
| | Hand | | 5.9% (0.1-28.7) | 13.0% (2.8-33.6) | 4.7% (1.3-11.6) |
| | Head | 4.4% (0.5-15.2) | | 8.7% (1.1-28.0) | 4.7% (1.3-11.6) |
| | Chest | 4.4% (0.5-15.2) | 5.9% (0.1-28.7) | | 3.5% (0.7-10.0) |
| | Foot | 4.4% (0.5-15.2) | | | 2.4% (0.3-8.2) |
| | Hip | 4.4% (0.5-15.2) | | | 2.4% (0.3-8.2) |
| | Back | 2.2% (0.1-11.8) | | | 1.2% (0.0-6.4) |
| | Eye | 2.2% (0.1-11.8) | | | 1.2% (0.0-6.4) |
| | Face | | | 4.3% (0.1-22.0) | 1.2% (0.0-6.4) |
| Cut | | **4.4% (0.5-15.2)** | **29.4% (10.3-56.0)** | **26.1% (10.2-48.4)** | **15.3% (8.4-24.7)** |
| | Hand | 2.2% (0.1-11.8) | 17.6% (3.8-43.4) | 26.1% (10.2-48.4) | 11.8% (5.8-20.6) |
| | Head | 2.2% (0.1-11.8) | 5.9% (0.1-28.7) | | 2.4% (0.3-8.2) |
| | Face | | 5.9% (0.1-28.7) | | 1.2% (0.0-6.4) |
| Musculoskeletal Injury (ie Strain) | | **15.6% (6.5-29.5)** | | **13.0% (2.8-33.6)** | **11.8% (5.8-20.6)** |
| | Back | 4.4% (0.5-15.2) | | 13.0% (2.8-33.6) | 5.9% (1.9-13.2) |
| | Shoulder | 6.7% (1.4-18.3) | | | 3.5% (0.7-10.0) |
| | Arm | 2.2% (0.1-11.8) | | | 1.2% (0.0-6.4) |
| | Leg | 2.2% (0.1-11.8) | | | 1.2% (0.0-6.4) |
| Fracture | | **13.3% (5.1-26.8)** | **11.8% (1.5-36.4)** | **4.3% (0.1-22.0)** | **10.6% (5.0-19.2)** |
| | Hand | 6.7% (1.4-18.3) | 11.8% (1.5-36.4) | | 5.9% (1.9-13.2) |
| | Leg | 4.4% (0.5-15.2) | | | 2.4% (0.3-8.2) |
| | Foot | 2.2% (0.1-11.8) | | 4.3% (0.1-22.0) | 2.4% (0.3-8.2) |
| Puncture wound | | | **29.4% (10.3-56.0)** | **4.3% (0.1-22.0)** | **7.1% (2.6-14.7)** |
| | Hand | | 23.5% (6.8-49.9) | 4.3% (0.1-22.0) | 5.9% (1.9-13.2) |
| | Arm | | 5.9% (0.1-28.7) | | 1.2% (0.0-6.4) |
| Scratch | | **2.2% (0.1-11.8)** | **11.8% (1.5-36.4)** | **4.3% (0.1-22.0)** | **4.7% (1.3-11.6)** |
| | Hand | | 11.8% (1.5-36.4) | | 2.4% (0.3-8.2) |
| | Head | 2.2% (0.1-11.8) | | 4.3% (0.1-22.0) | 2.4% (0.3-8.2) |
| Crush injury | | **4.4% (0.5-15.2)** | | | **2.4% (0.3-8.2)** |
| | Back | 2.2% (0.1-11.8) | | | 1.2% (0.0-6.4) |
| | Foot | 2.2% (0.1-11.8) | | | 1.2% (0.0-6.4) |
| Dislocation | | **4.4% (0.5-15.2)** | | | **2.4% (0.3-8.2)** |
| | Shoulder | 2.2% (0.1-11.8) | | | 1.2% (0.0-6.4) |
| | Teeth | 2.2% (0.1-11.8) | | | 1.2% (0.0-6.4) |
| Allergic Response | | | | **4.3% (0.1-22.0)** | **1.2% (0.0-6.4)** |
| | Head | | | 4.3% (0.1-22.0) | 1.2% (0.0-6.4) |

| | | | | | |
|---|---|---|---|---|---|
| Burn | | | | **4.3% (0.1-22.0)** | **1.2% (0.0-6.4)** |
| | Hand | | | 4.3% (0.1-22.0) | 1.2% (0.0-6.4) |
| Chemical Irritation | | | | **4.3% (0.1-22.0)** | **1.2% (0.0-6.4)** |
| | Eye | | | 4.3% (0.1-22.0) | 1.2% (0.0-6.4) |
| Concussion | | **2.2% (0.1-11.8)** | | | **1.2% (0.0-6.4)** |
| Loss of consciousness | | **2.2% (0.1-11.8)** | | | **1.2% (0.0-6.4)** |
